# Effect of multiple freeze-thaw cycles on detection of anti-SARS-CoV-2 IgG antibodies

**DOI:** 10.1101/2021.04.13.21255379

**Authors:** Farah M. Shurrab, Duaa W. Al-Sadeq, Fathima Amanullah, Salma N. Younes, Hadeel Al-Jighefee, Nadin Younes, Soha R. Dargham, Hadi M. Yassine, Laith J. Abu Raddad, Gheyath K. Nasrallah

**Affiliations:** Biomedical Research Center, Qatar University, Doha 2713, Qatar; College of Medicine, Member of QU Health, Qatar University, P.O. Box 2713, Doha, Qatar; Department of Biomedical Science, College of Health Sciences, Member of QU Health, Qatar University, P.O. Box 2713, Doha, Qatar; Infectious Disease Epidemiology Group, Weill Cornell Medicine□Qatar, Cornell University, Qatar Foundation – Education City, Doha, Qatar; World Health Organization Collaborating Centre for Disease Epidemiology Analytics on HIV/AIDS, Sexually Transmitted Infections, and Viral Hepatitis, Weill Cornell Medicine–Qatar, Cornell University, Qatar Foundation – Education City, Doha, Qatar; Department of Healthcare Policy and Research, Weill Cornell Medicine, Cornell University, New York, United States of America

**Keywords:** Antibodies, Detection, COVID-19, SARS-CoV-2, IgG, Freeze, Thaw

## Abstract

Several studies have investigated the effect of repeated freeze-thaw (F/T) cycles on RNA detection for SARS-CoV-2. However, no data is available regarding the effect of repeated F/T cycles on SARS-CoV-2 antibody detection in serum. We investigated the effect of multiple F/T cycles on anti-SARS-CoV-2 IgG detection using an ELISA test targeting the nucleocapsid antibodies. Ten positive and one negative SARS-CoV-2 IgG sera from 11 participants, in replicates of five were subjected to a total of 16 F/T cycles and stored at 4°C until tested by ELISA. Statistical analysis was done to test for F/T cycle effect. Non-of the 10 positive sera turned into negative after 16 F/T cycles. There was no significant difference in the OD average reading between the first and last F/T cycles, except for one serum with a minimal decline in the OD. The random-effect linear regression of log (OD) on the number of cycles showed no significant trend with a slope consistent with zero (B=-0.0001; 95% CI −0.0008; 0.0006; p-value=0.781). These results suggest that multiple F/T cycles had no effect on the ability of the ELISA assay to detect the SARS-CoV-2 IgG antibodies.

## 1. Introduction

Serum banks are well-established as they are considered an essential reference for clinical information and research use. However, there are concerns regarding the effect of repeated freeze-thaw (F/T) cycles on the biological entities of serum proteins, including immunoglobulins (Ig) (1-4). It is suspected that repeated F/T cycles may lead to denaturation or degradation of the antibody of interest (5). This is critical when it comes to sensitive immunoassays such as ELISA or chemiluminescence automated analyzers that detect antibodies in serum or plasma. Therefore, it is recommended to store sera in aliquots to reduce sample exposure to multiple F/T cycles (6). Although several studies suggested that repeated F/T cycles have a minimal effect on antibody stability against specific pathogens (2, 7, 8), other researchers are questioning the reliability of the data generated from using such samples (9).

The emergence of the severe acute respiratory syndrome coronavirus type 2 (SARS-CoV-2) in late December 2019 in Wuhan, China, has led to a global Coronavirus Disease 2019 (COVID-19) pandemic (10). Several studies showed the effect of repeated F/T cycles on SARS-CoV-2 RNA stability in the throat and nasopharyngeal swabs specimens (11, 12). However, to the best of our knowledge, the stability of SARS-CoV-2 antibodies after multiple F/T cycles has not been assessed. In this study, we investigated the effect of multiple F/T cycles on SARS-CoV-2 IgG detection in serum by using ELISA targeting the nucleocapsid (N) antibodies.

## 2. Materials and Methods

The cohort sera used in this study were part of blood specimens that were collected in a previous nationwide survey to assess prevalence of SARS-CoV-2 detectable antibodies (13). The specimens were collected between July 26 and September 9, 2020 and frozen and thawed twice during the previous study before being transferred on ice to our facility at Qatar University, where they were stored once more at −80 C° until used in this study, in December of 2020.

Fifty sera were screened using the EDI™ novel coronavirus COVID-19 IgG ELISA kit (Ref. no. KT-1032, USA) targeting the anti-N SARS-CoV2 IgG (14). Eleven sera were selected of these; of which, 10 were IgG positive and one was IgG negative. The latter serum was used as a control. For a more representative comparison between measurements, we selected the positive sera that had broadly different optical density (OD) readings (high, medium, and low).

From each of the eleven sera, a total of 40 serum aliquots, 5µl each, were divided into 8 sets of five replicates and subjected to 8 different freeze-thaw cycles (3, 4, 6, 8, 10, 12, 14, and 16). The first set was stored at 4°C during the study as a baseline. The remaining 7 sets were subjected to the repeated freeze-thaw cycles, with one set of aliquots being stored at 4°C at a time until all cycles were completed. The serum samples were then tested by the EDI kit and the OD reading at 450 nm was recorded.

The average reading of each serum was estimated and plotted against the number of F/T cycles and versus the cut-off values defining a positive or a negative outcome. The cut offs values were calculated according to the manufacturer instructions. Independent t-test was conducted to compare the OD measurements of the positive sera and the negative serum. Paired t-test were performed to compare the log (OD) of the first cycle to the log (OD) of the last cycle. To adjust for any clustering effect from measurements of the same serum, a random-effect linear regression was conducted on the log (OD) versus the number of F/T cycles. A p-value of <0.05 was considered statistically significant.

## 3. Results

A total of 438 measurements from 11 sera were available for analysis. Two OD readings were excluded due to manual error during ELISA run. The mean OD values for the positive serum measurements [N=398; mean (SD): 0.69 (0.25)] was significantly higher than that of the negative serum measurement [N=40; mean (SD): 0.23 (0.03); p-value<0.001].

The average assay readings for each serum were plotted against the F/T cycles and versus the cut-off values delineated at ≤0.2655 for negatives and ≥0.3245 for positives (Figure 1). When antibody OD readings were analyzed as categorical outcomes (positive, negative, and equivocal) across the cycles, out of 398 positive measurements, 0.8% (3/398; 95% CI 0.3%-2.2%) were no longer classified as positive using the positive cut-off. Instead, there were equivocal. Out of 40 negative measurements, 20.0% (8/40; 95% CI 10.5%-34.8%) were no longer classified as negative using the negative cut-off (Figure 2). Instead, there were equivocal. None of the sera changed from positive to negative or from negative to positive throughout the cycles.

**Figure 1.**
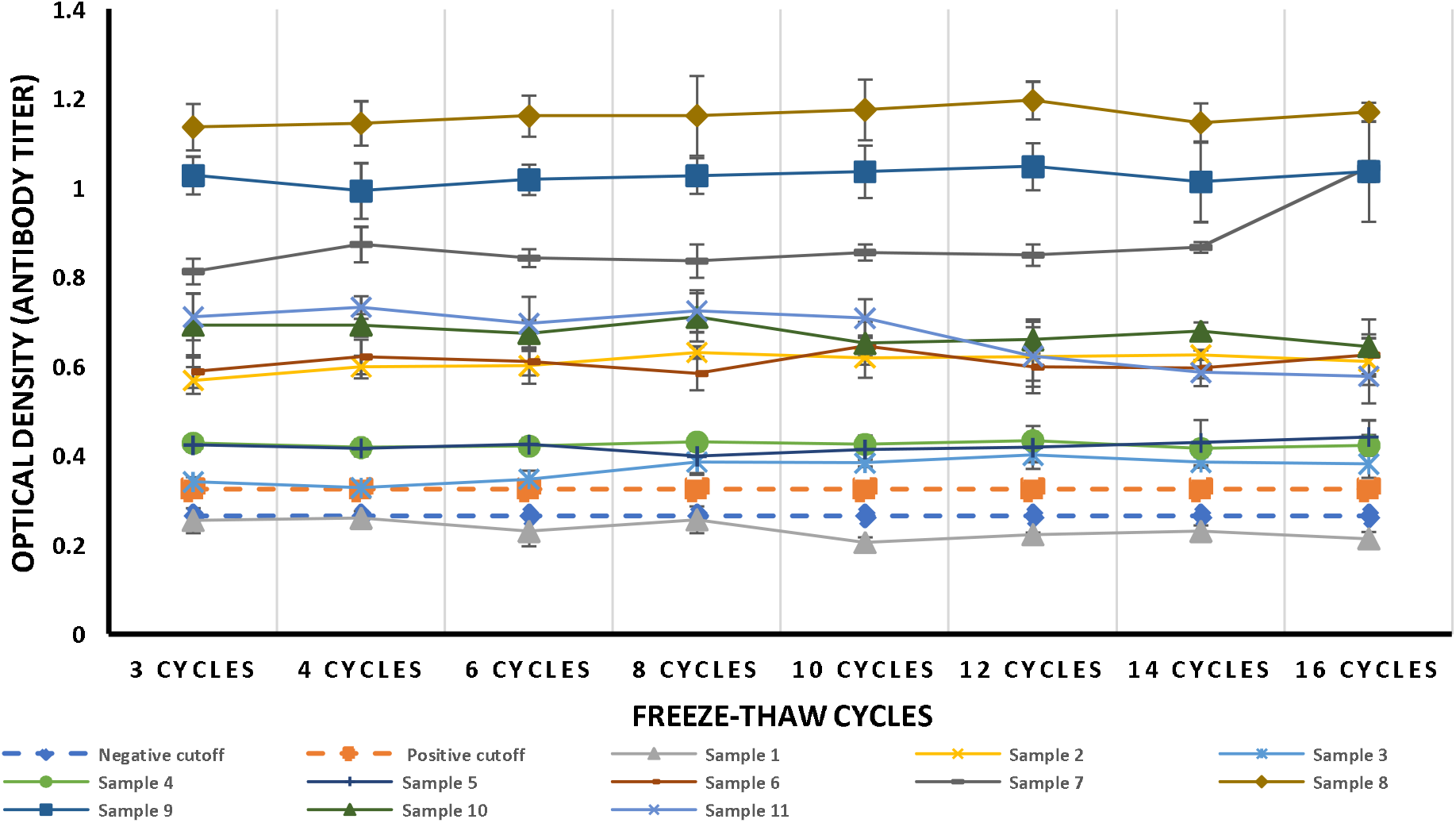
SARS-CoV-2 IgG antibody average 450 optical density readings of the 5 replicates for each serum plotted against the number of freeze-thaw cycles.

**Figure 2.**
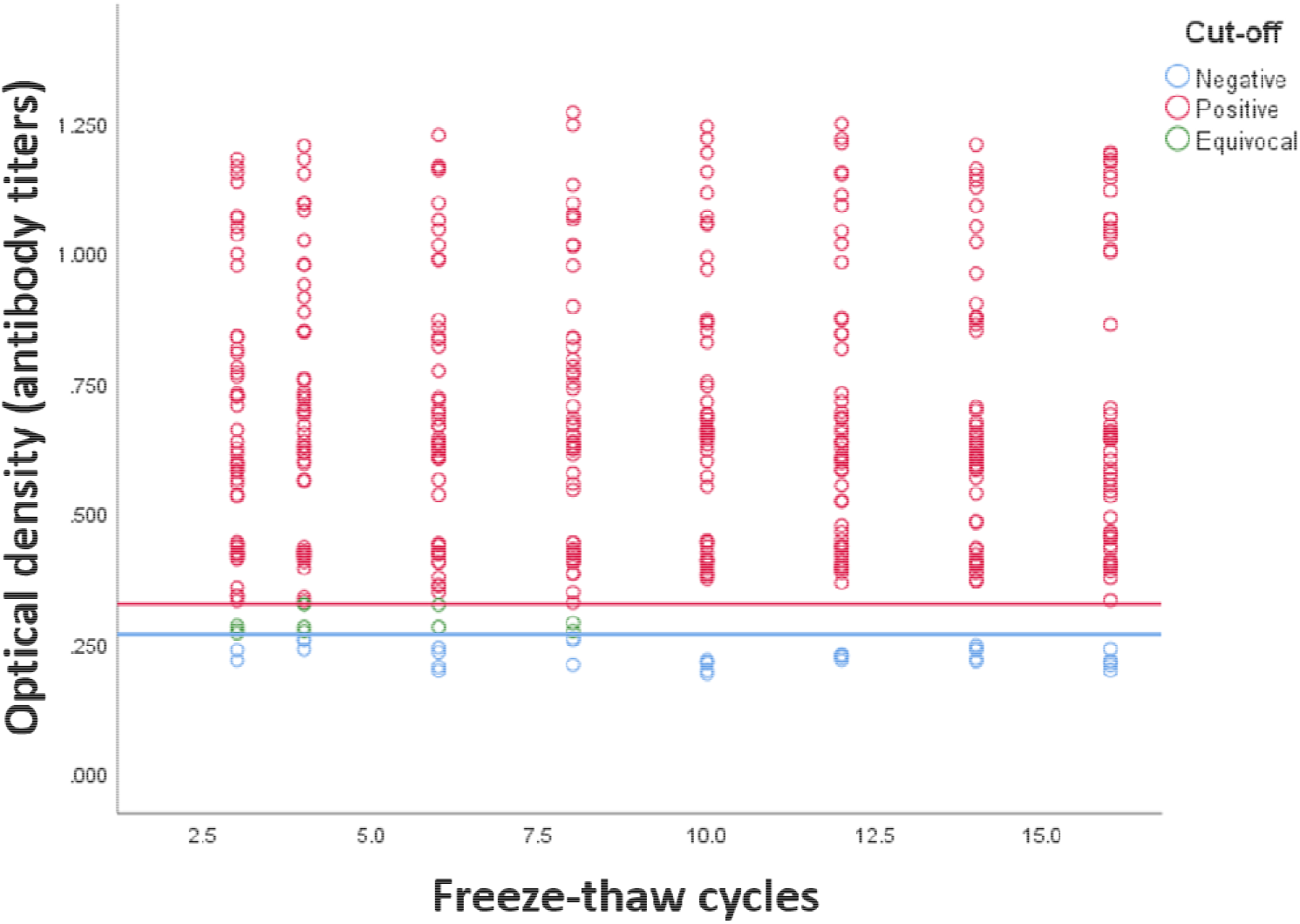
Antibody levels for SARS-CoV-2 for 438 specimens against the number of freeze-thaw cycles. The blue and red solid horizontal lines delineate the cut-off for negative at optical density 0.2655 and the cut-off for positive at 0.3245.

The random-effect linear regression of log (OD) on number of cycles showed no significant trend with a slope consistent with zero (B=-0.0001; 95% CI −0.0008; 0.0006; P-value=0.781), indicating that the multiple F/T cycles had no effect on the SARS-CoV-2 IgG antibody titers in the serum.

## 4. Discussion

Frozen serum banks are an important source of scientific and clinical information and are essential to infectious disease and vaccine research. It is commonly believed that antibodies keep their stability if serum is stored below −20C° (9). Nevertheless, evidence suggests that just one F/T cycle can significantly reduce IgG antibody levels (9). To date, studies investigating the effects of F/T cycles on the detection of SARS-CoV-2 antibodies have yielded inconsistent results, and the data availability on the effects of multiple F/T cycles, sample size, and storage conditions, is limiting.

The present study investigated the effect of 16 repeated F/T cycles on SARS-CoV-2 IgG antibodies in serum. The results showed that in nearly all sera there was no significant difference in sample reactivity between the first and last F/T cycle, and none of the reactive sera became non-reactive after 16 F/T cycles, or any false-positive results were obtained (Figure 2). The random effect linear regression showed no significant trend in sera OD reading with a slope that is consistent with zero. However, there was one outlier with one serum showing an increased OD reading after cycle 16 (Figure 1, sample no. 7). This outlier may have been caused by a pipetting error between the ELISA plates.

In agreement with our findings, other studies have tested the effect of repeated F/T cycles on measles, mumps, rubella, syphilis, anti-nuclear antibodies (ANA), and anti-neutrophil cytoplasmic antibodies (ANCA), and found similar results (2, 8, 15). Although each study applied a different freeze-thaw methodology and targeted different antigens, they all concluded that there was no clinically or statistically significant difference in the antibody levels after several F/T cycles. Interestingly, another study has shown that 174 F/T cycles on anti-treponemal sera did not affect the stability, the reactivity of antibodies, nor the samples’ quality when tested by a chemiluminescence assay (1). The data generated from our study and the previous studies provide concrete knowledge regarding antibody stability in serum, which allows the maximum potential use of serum, especially for those that have undergone multiple F/T cycles. Additionally, our generated results can be beneficial to the design of serum banks, where it is important to monitor the integrity of sample components.

We conclude that 16 F/T cycles did not interfere with the detection of SARS-CoV-2 IgG antibodies, and with no effect on assay sensitivity. However, in this study, we tested the effect of multiple F/T cycles on anti-SARS-CoV-2 IgG in serum only. It would be ideal to use plasma samples as well and to test the stability of other anti-SARS-CoV-2 antibody classes such as IgM and IgA. Furthermore, our ELISA test targeted antibodies against SARS-CoV-2 nucleocapsid protein alone, but possibly F/T cycles may interfere with other SARS-CoV-2 proteins, such as the spike protein. Lastly, using an automated serological analyzer would be beneficial to reduce pipetting errors.

## Data Availability

We were constrained by our local, internal ethics policies, which entail confidentiality and therefore anonymity with regard to the samples for analysis in studies of this nature. Disclosing raw data will be breaching participant confidentiality requests to access the data should be addressed to the Ethical committee of Qatar University

## Author contribution

Conceptualization: GKN, HMY, LJA; Methodology: FMS, HAJ, DWA, SNY, FHA. Formal Analysis: SRD, LJA, NY; Validation: GKN, FMS; Investigation: FMS, GKN, SRD, LJA, Resources: GKN, HMA; Data Curation: GKN, FMS; Writing – Original Draft Preparation: FMS, GKN, SRD, LJA; Writing, Review & Editing: DWA, HAJ, SRD, GKN, HMY, LJA; Visualization: GKN, FMS; Supervision: GKN, FMS; Project Administration: GKN, DWA, Funding Acquisition: GKN, HMY, LJA. All authors have read and agreed to the published version of the manuscript.

## Acknowledgements

We would like to thank Dr. Nahla Afifi, Director of Qatar Biobank (QBB), Ms. Tasneem Al-Hamad, Ms. Eiman Al-Khayat and the rest of the QBB team for their unwavering support in retrieving and analyzing samples and in compiling and generating databases for COVID-19 infection, as well as Dr. Asma Al-Thani, Chairperson of the Qatar Genome Programme Committee and Board Vice Chairperson of QBB, for her leadership of this effort.

## Ethical statement

This study was approved by the Medical Research Center (MRC) Ethics Committee, approval number MRC-05-136, and Qatar University Institutional Review Board approval number QU-IRB 1492-E/21.

## References

1. Castejon MJ, Yamashiro R, Oliveira CC, Oliveira EL, Silveira EP, Oliveira CAFJJBdPeML. Effect of multiple freeze-thaw cycles on the stability of positive anti-treponemal serum samples. 2017;53(4):246–51.

2. Castro AR, Jost HAJStd. Effect of multiple freeze and thaw cycles on the sensitivity of IgG and IgM immunoglobulins in the sera of patients with syphilis. 2013;40(11):870–1.

3. Guo GH, Dong J, Yuan XH, Dong ZN, Tian YPJMmr. Clinical evaluation of the levels of 12 cytokines in serum/plasma under various storage conditions using evidence biochip arrays. 2013;7(3):775–80.

4. Cuhadar S, Koseoglu M, Atay A, Dirican AJBmBm. The effect of storage time and freeze-thaw cycles on the stability of serum samples. 2013;23(1):70–7.

5. Miller MA, Rodrigues MA, Glass MA, Singh SK, Johnston KP, Maynard JAJJops. Frozen-state storage stability of a monoclonal antibody: aggregation is impacted by freezing rate and solute distribution. 2013;102(4):1194–208.

6. Paltiel L, Rønningen KS, Meltzer HM, Baker SV, Hoppin JAJCpt. Evaluation of freeze–thaw cycles on stored plasma in the biobank of the Norwegian Mother and Child Cohort study. 2008;6(3):223–9.

7. Maelegheer K, Devreese KMJR, thrombosis pi, haemostasis. The impact of repeated freeze-thaw cycles on antiphospholipid antibody titer. 2018;2(2):366–9.

8. Pinsky NA, Huddleston JM, Jacobson RM, Wollan PC, Poland GAJC, immunology dl. Effect of multiple freeze-thaw cycles on detection of measles, mumps, and rubella virus antibodies. 2003;10(1):19–21.

9. Petrakis NL. Biologic banking in cohort studies, with special reference to blood. National Cancer Institute monograph. 1985;67:193–8.

10. Younes S, Younes N, Shurrab F, Nasrallah GKJRimv. Severe acute respiratory syndrome coronavirus-2 natural animal reservoirs and experimental models: systematic review. 2020:e2196.

11. Li L, Li X, Guo Z, Wang Z, Zhang K, Li C, et al. Influence of storage conditions on SARS-CoV-2 nucleic acid detection in throat swabs. 2020.

12. Stohr JJ, Wennekes M, van der Ent M, Diederen BM, Kluytmans-van den Bergh MF, Bergmans AM, et al. Clinical performance and sample freeze-thaw stability of the cobas^®^ 6800 SARS-CoV-2 assay for the detection of SARS-CoV-2 in oro-/nasopharyngeal swabs and lower respiratory specimens. 2020:104686.

13. Al-Thani MH, Farag E, Bertollini R, Al Romaihi HE, Abdeen S, Abdelkarim A, et al. Seroprevalence of SARS-CoV-2 infection in the craft and manual worker population of Qatar. medRxiv. 2020:2020.11.24.20237719.

14. Yassine HM, Al-Jighefee H, Al-Sadeq DW, Dargham SR, Younes SN, Shurrab F, et al. Performance evaluation of five ELISA kits for detecting anti-SARS-COV-2 IgG antibodies. 2020;102:181–7.

15. Demir M, Cevahir N. Does multiple freezing and thawing cycles of serum affect the detection of anti-nuclear antibodies and anti-neutrophil cytoplasmic antibodies by indirect immunofluorescent method? 2014.

